# Impact of the Federated Data Platform’s digital surgery scheduling system on elective theatre utilisation at an NHS Trust: an interrupted time series analysis

**DOI:** 10.1101/2025.11.24.25340393

**Authors:** Elena Lammila-Escalera, Gabriele Kerr, Geva Greenfield, Benedict Hayhoe, Natalie Brewer, Carla Hearsum, Grazia Antonacci, Natasha Dsouza, Azeem Majeed, Ana Luisa Neves

**Affiliations:** Department of Primary Care and Public Health, Imperial College London; Department of Primary Care and Public Health; Digital Operations and Innovation, Chelsea and Westminster Hospital NHS Foundation Trust; Chelsea and Westminster Hospital NHS Foundation Trust Lead for Theatres, Chelsea and Westminster Hospital NHS Foundation Trust; Deputy Lead for Innovation and Evaluation Theme at NIHR Applied Research Collaboration Northwest London and Research Fellow, Department of Primary Care and Public Health and the Centre for Health Economics and Policy Innovation, Imperial College London; Innovation and Evaluation Theme at NIHR Applied Research Collaboration Northwest London, Department of Primary Care and Public Health, Imperial College London; Public Health and Head of the Department of Primary Care and Public Health, Department of Primary Care and Public Health, Imperial College London; Digital Health and Director of the Global Digital Health Unit, Department of Primary Care and Public Health, Imperial College London

## Abstract

**Objectives:** To evaluate the NHS Federated Data Platform (FDP) Inpatient Care Coordination Solution (CCS) digital scheduling tool on elective theatre utilisation.

**Methods:** An interrupted time series assessed changes in theatre utilisation and cancellations following tool adoption (January 2022). Weekly data spanned 90 weeks (April 2021 to December 2023). Outcomes included weekly median theatre utilisation (actual, booked, and bookings per session) and the percentage of cancelled bookings. Models incorporated a 5-week lag and estimated level (step-change) and trend (slope) effects.

**Results:** Post-intervention level and trend increases were observed for booked (β=4.40*, P*=0.045; β=0.26*, P*=0.002) and actual (β=3.98*, P*=0.064; β=0.23*, P*=0.006) utilisation. Bookings per session showed a significant level increase (β=0.34*, P*=0.002) with no trend change (β=0.00*, P*=0.790). Across the post-intervention period, compared with counterfactual estimates, booked and actual utilisation were 15.0% (95% CI: 13.4 to 16.5%, *P*<0.0001) and 12.2% (95% CI: 10.8 to 13.5%, *P*<0.0001) higher, while bookings per session were 10.9% (95% CI: 9.5 to 12.4%, *P*<0.0001) higher. Significant positive effects were observed for Urology, General Surgery, Gynaecology, Plastic Surgery and Ophthalmology. Cancellation rates remained stable pre- and post-intervention (5.2% vs. 5.2%, *P=*0.488), but a significant upward trend was associated with the tool (β=2.1, *P*=0.001).

**Discussion:** Findings suggest that a centralised digital scheduling tool can improve theatre capacity, while a small increase in cancellation trend likely reflects post-pandemic recovery or improved recording. Future research should explore speciality-level use and the sustainability of observed gains.

**Conclusion:** The introduction of the CCS tool was associated with improved theatre utilisation.

**Summary box:** *What is already known on the topic:*

- Inefficient surgical scheduling results in delays in care, suboptimal patient outcomes, and increased costs for healthcare providers.
- Digital theatre scheduling systems have been developed to enhance scheduling efficiency; however, evidence of their effectiveness from real-world implementations remains limited.
- Most previous studies focus on theoretical optimisation models rather than practical, system-wide impact in routine care settings.

*What this study adds:*

- Provides real-world evidence that integrated digital scheduling platforms can enhance elective theatre scheduling, indicating a more efficient use of existing capacity despite variation across specialities.

*How this study might affect research, practice or policy:*

- Enhanced scheduling efficiency benefits patients by minimising wait times and reducing the risks associated with delays, strengthening the argument for adopting such tools to address the current backlog in elective care.
- Identifies priorities for future research, including long-term evaluation, factors influencing adoption, and the role of digital scheduling in elective recovery.

## Introduction

Operating theatres are among the most resource-intensive areas of secondary care.[1,2] Consequently, maximising theatre utilisation is critical in controlling care costs.[3] An NHS Improvement audit in 2019 found that 38% of theatre lists were underutilised, with unused theatre time estimated to cost the NHS approximately £400 million annually.[3,4] In 2017 alone, time lost was equivalent to 27,000 oral and maxillofacial surgical procedures.[3]

Poor utilisation reduces surgical capacity, contributing to same-day cancellations, treatment delays, and pressure on downstream services.[5–7] As of August 2025, only 53.8% of patients began treatment within 18 weeks, which is significantly below the 92% target.[8] Over 191,500 patients were waiting more than a year to start care.[8–10] Such prolonged delays negatively impact clinical outcomes, including higher mortality risk, clinical deterioration and increased treatment complexity. [11–13] They also have a substantial impact on patients’ quality of life, contributing to pain and loss of independence,[14–16] and are frequently accompanied by frustration, social isolation and depressive symptoms.[14,17] Notably, one in three patients whose surgeries are cancelled report being unable to carry out daily activities while awaiting rescheduling.[18]

An important determinant of theatre utilisation is the efficiency of surgery scheduling.[19] Elective procedures frequently compete with emergency cases, while unpredictable operation times can disrupt even well-organised schedules, making list reconstruction time-consuming and inefficient.[20–22] In recent years, improving theatre productivity has become a central priority for NHS elective care reform, with a range of interventions introduced to address these challenges.[23]

Digital systems present new opportunities to improve surgical scheduling and theatre management through more accurate forecasting, dynamic resource allocation, and data-driven decision-making. [24] Emerging platforms leverage advanced analytics and real-time data to create adaptive schedules, enhance visibility, identify bottlenecks, and facilitate coordinated team planning.[20] However, evidence on their real-world impact remains limited, as most existing studies focus on operational scheduling or optimisation methods, rather than practical implementation and real-world implications.[25–33]

One such solution is the NHS Federated Data Platform’s (FDP) Inpatient (IP) Care Coordination Solution (CCS) product theatre management tool, originally developed at Chelsea and Westminster NHS Foundation Trust (CWFT), refined through pilot use at several other NHS trusts. The module is now widely accessible to NHS acute trusts via NHSE’s FDP Programme and is presently in use at 39 trusts.[34] Designed to optimise elective surgery scheduling, the system features prominently in NHS digital transformation strategies but remains under-evaluated. This study addresses this gap by assessing the impact of the FDP IP theatre management tool on a) theatre utilisation (i.e. booked utilisation, actual utilisation, bookings per theatre session) and b) cancellation rates at CWFT.

## Methods

### Setting

CWFT consists of two hospitals (Chelsea and Westminster Hospital and West Middlesex University Hospital) based in Northwest London, serving a population of 1.5 million people and employing 7,500 staff members.[35] This study adheres to the STROBE guidelines for reporting observational studies.[36]

### Intervention

The intervention is the NHS FDP Inpatient CCS product. The NHS FDP is a national digital infrastructure initiative that provides each organisation with its own dedicated data platform, enabling trusts to connect and share information stored in separate systems. This allows staff to access the information required in a single, secure, and safe environment.[37–39] The FDP Inpatient CCS product theatre management module, one of several modules within the FDP ecosystem, is currently deployed by 32 NHS trusts.[40] CWFT implemented the FDP Inpatient CCS product across their two sites in January 2022 to optimise elective surgery scheduling and waiting list management. The intervention was introduced at CWFT on January 3^rd^, 2022. The tool provides a centralised view of theatre sessions and lists, enabling users (staff or schedulers) to identify available sessions, cancel or reassign bookings, and ensure theatres are booked to capacity **(Supplementary Appendix, Pages 2 and Figures S1-5**).

### Data sources

To reflect the scheduling cycles of surgical staff, a five-week lag was applied, yielding a modelled intervention start of 6^th^ February 2022. All data were accessed through the NHS FDP secure environment. Data tables related to theatre bookings (1^st^ January 2018 to 1^st^ January 2025) and cancelled cases (7^th^ of May 2018 to 29^th^ of April 2025) were extracted.

The analytical period was restricted to 28^th^ March 2021 and 17^th^ December 2022 to minimise COVID-19-related service disruption and to ensure a stable baseline for post-pandemic elective activity. This start followed a sharp decline in utilisation in January 2021 due to suspended elective surgery, temporary site closures, ward consolidation and staff redeployment during the second national lockdown.[41] By late March 2021, targeted recruitment had stabilised staffing levels and surgical rotas, providing a consistent baseline for this study, despite disruption continuing throughout 2021.[42,43]

### Outcomes

A hybrid workshop was held on 19 July 2024 to develop the logic model and identify key metrics for evaluating the FDP Inpatient CCS theatre management platform with relevant stakeholders. Seven subject matter experts from three NHS trusts (The Hillingdon Hospital NHS Foundation Trust, Imperial College Healthcare Trust, and London North West University Healthcare NHS Trust) participated. The session aimed to establish a shared understanding of the FDP IP platform’s theatre session management functions and relevant workflows (**Supplementary Appendix Page 8**). The research team facilitated and conducted a thematic analysis of the workshop findings, which informed the selection of metrics (**Supplementary Appendix Figure S6**).

Additionally, complementary materials (i.e., online training materials and standard operating procedures, incorporating workflow maps and stakeholder guides for managers, schedulers, clinicians, and theatre staff) were reviewed to inform the development of a logic model [44], describing causal pathways between activities and outcomes, and the final evaluation metrics (**Table 1**).

**Table 1:** Key metrics and their definitions.

| Type of metric | Metric | Definition |
| --- | --- | --- |
| Theatre utilisation | Actual utilisation | Actual session utilisation ratio, defined as (actual utilised time / available time) * 100 |
|  | Booked utilisation | Booked utilisation ratio, defined as (minutes booked / total minutes available) * 100 |
|  | Number of bookings per theatre session | Count of booked patients, not including the number of booked patients in theatre sessions that were cancelled or rescheduled. |
| Cancellations | Cancellation rate | Cancellation rate defined as the proportion of bookings that were cancelled in a given period (number of cancelled bookings / total number of bookings). |

### Statistical Analysis

A quasi-experimental interrupted time series (ITS) design was used to evaluate the impact of the intervention. Outcome data were input as weekly averages, yielding 45 weeks of pre-intervention training data; the study period end was therefore capped at 45 weeks post-intervention to maintain balance.[45]

ITS models estimated both level (i.e., immediate effects or step changes) and trend (longer-term changes in slope) changes. Outcomes were modelled using segmented generalised least squares regression.[45,46] A linear time-outcome relationship was assumed within each segment. The ITS design assumed: 1) a single intervention adoption point across the Trust; 2) no concurrent interventions affecting utilisation; 3) consistent FDP Inpatient CCS use post-intervention; and 4) absence of unobserved time-varying confounders (e.g., significant changes in the patient population). Models for booked and actual utilisation incorporated a first-order autoregressive structure based on autocorrelation diagnostics, while cancellation rates were log-transformed to meet normality assumptions. Seasonality in utilisation models was adjusted using Fourier terms. The full model development process is described in **Supplementary Appendix Page 9**. Model residuals were inspected for normality, zero mean, and autocorrelation, with the diagnostic results presented in **Supplementary Appendix Figures S7-10**.

For each outcome, the model estimated the underlying trend and a hypothetical counterfactual (no intervention) trajectory. To compare observed and counterfactual means, 10,000 simulations of the counterfactual scenario were generated by sampling at each time point from a normal distribution defined by the predicted mean and its standard error. For each simulated dataset, the mean difference between observed and counterfactual predicted values was calculated and expressed as a percentage of the mean counterfactual value. The distribution of the simulated percentage differences was then summarised to obtain the posterior mean and 95% credible interval.

Analyses were performed for the overall Trust and stratified by surgical speciality; only specialities averaging at least three theatre sessions per week were included in the stratified analyses. All analyses were conducted in R software version 4.3.0.[47], with a statistical significance threshold of α <0.05 used throughout.

### Patient and public involvement

There was no patient or public involvement in the development or execution of this study. The results of this study will be shared through organisational patient and public engagement events.

## Results

Between 28th March 2021 and 17th December 2022, there were 8,637 theatre sessions and 31,572 bookings, of which 1,883 (5.56%) were cancelled. The median booked and actual utilisation, the bookings per session, and the proportion of cancelled cases are summarised in **Table 2**. Utilisation metrics and cancellation rates over time are presented in **Supplementary Figures S11-13**.

**Table 2:**
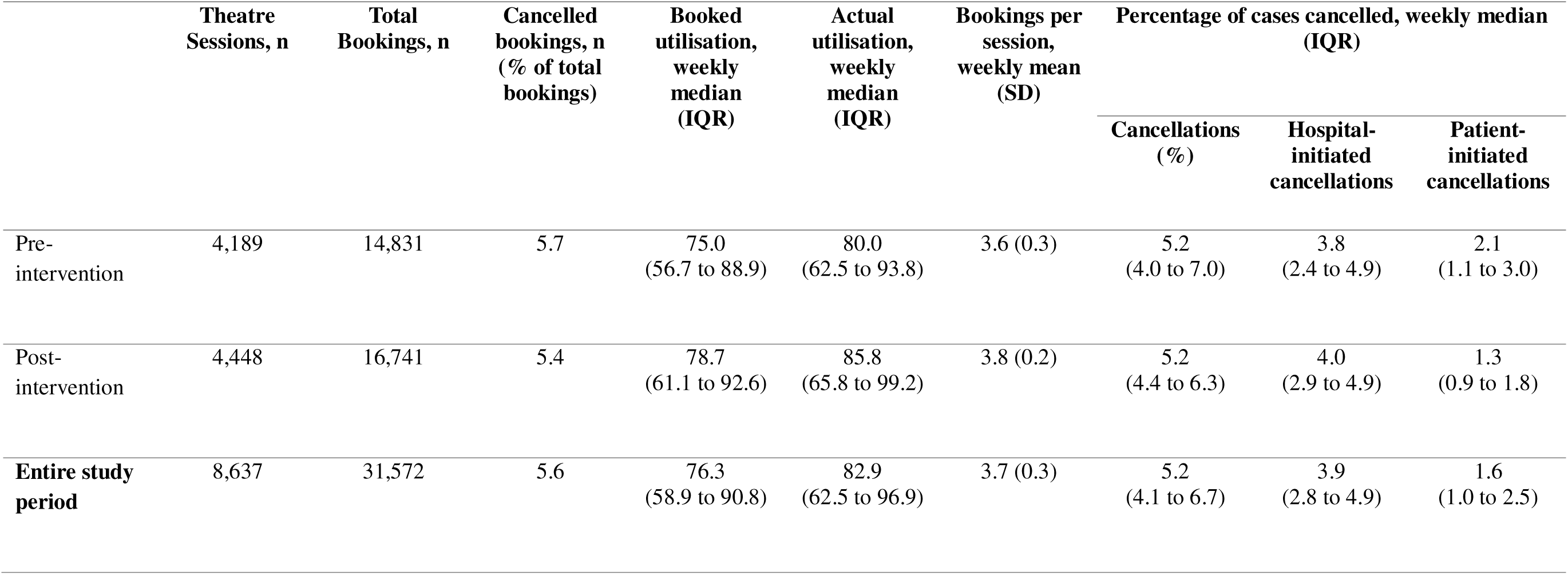
Changes in the values of key performance metrics before and after FDP Inpatient CCS adoption.

|  | Theatre Sessions, n | Total Bookings, n | Cancelled bookings, n (% of total bookings) | Booked utilisation, weekly median (IQR) | Actual utilisation, weekly median (IQR) | Bookings per session, weekly mean (SD) | Percentage of cases cancelled, weekly median (IQR) |  |  |
| --- | --- | --- | --- | --- | --- | --- | --- | --- | --- |
|  |  |  |  |  |  |  | Cancellations (%) | Hospital-initiated cancellations | Patient-initiated cancellations |
| Pre-intervention | 4,189 | 14,831 | 5.7 | 75.0<br>(56.7 to 88.9) | 80.0<br>(62.5 to 93.8) | 3.6 (0.3) | 5.2<br>(4.0 to 7.0) | 3.8<br>(2.4 to 4.9) | 2.1<br>(1.1 to 3.0) |
| Post-intervention | 4,448 | 16,741 | 5.4 | 78.7<br>(61.1 to 92.6) | 85.8<br>(65.8 to 99.2) | 3.8 (0.2) | 5.2<br>(4.4 to 6.3) | 4.0<br>(2.9 to 4.9) | 1.3<br>(0.9 to 1.8) |
| **Entire study period** | 8,637 | 31,572 | 5.6 | 76.3<br>(58.9 to 90.8) | 82.9<br>(62.5 to 96.9) | 3.7 (0.3) | 5.2<br>(4.1 to 6.7) | 3.9<br>(2.8 to 4.9) | 1.6<br>(1.0 to 2.5) |

### Theatre Utilisation and Bookings per Session

An increasing trend was observed in median weekly booked utilisation following the intervention (**Table 3**, **Figure 1A**). By six months after the implementation of the tool, booked utilisation increased to 84.3%, compared with an estimated 71.2% (95% CI: 65.2 to 77.2%) if the tool had not been introduced. Mean booked utilisation for the whole post-intervention study period was 15.0% (95% CI 13.4 to 16.5%, p < 0.0001) higher than expected compared to the counterfactual estimate.

**Figure 1:**
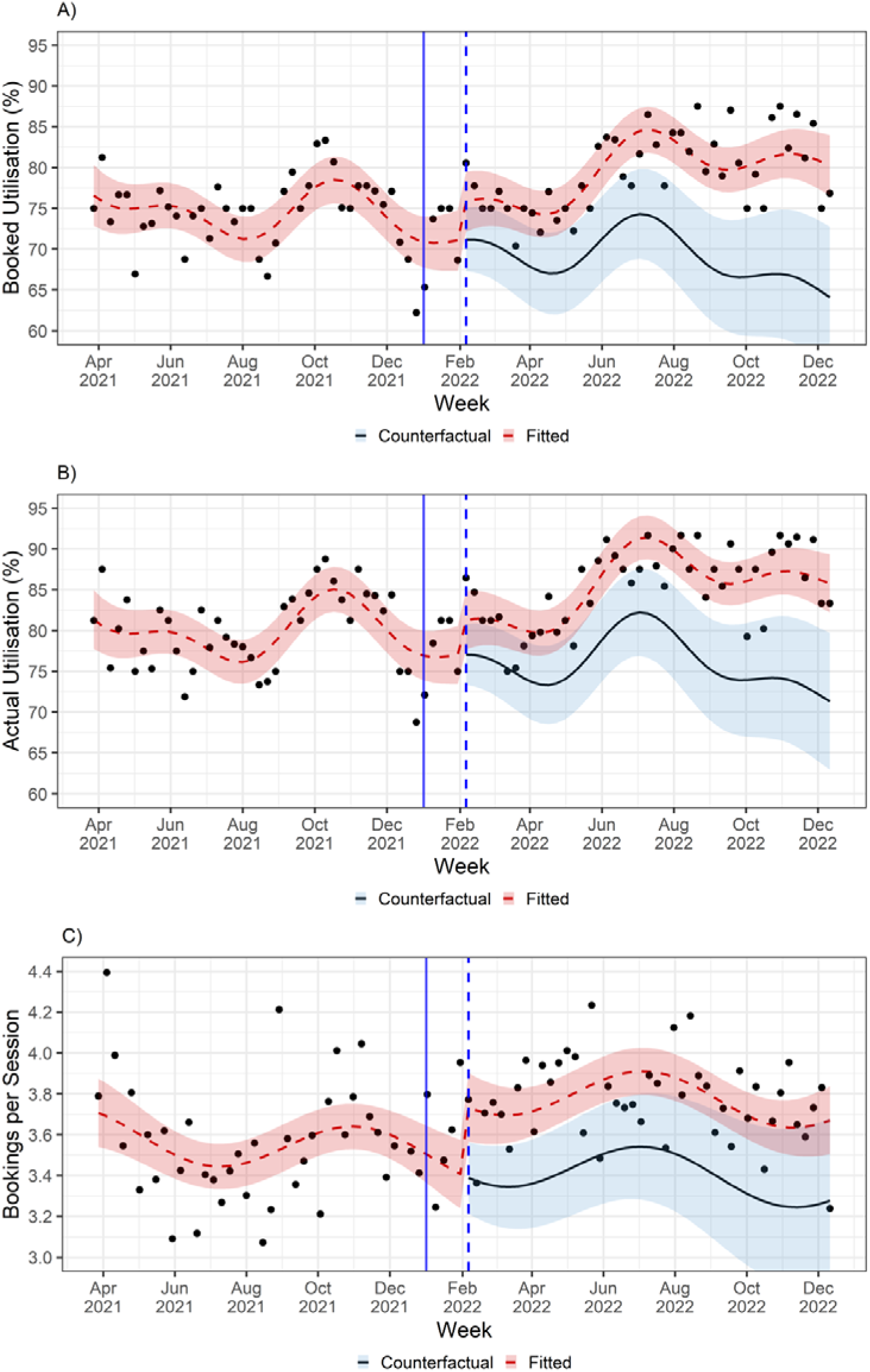
Changes in weekly a) median booked theatre utilisation, b) median actual theatre utilisation, and c) number of bookings per theatre session, following introduction of the FDP Inpatient CCS product on January 3^rd^, 2022. Models assumed a five-week lag in start of the effect of the intervention. The blue dashed line indicating the modelled five-week lag in the start-date. The red line shows the modelled fit to the observed results. The black line represents the counterfactual scenario (i.e., the expected results if there had been no intervention). The shaded areas around each line represent 95% confidence intervals.

**Table 3:** Results from interrupted time series analyses of median weekly booked and actual utilisation, and weekly bookings per session.

|  | Booked utilisation |  | Actual utilisation |  | Bookings Per Session |  |
| --- | --- | --- | --- | --- | --- | --- |
|  | Value<br>(95% CI) | p-value | Value<br>(95% CI) | p-value | Value (95% CI) | p-value |
| **Level impact** | 4.40<br>(0.10 to 8.70) | 0.045 | 3.98<br>(-0.18 to 8.10) | 0.064 | 0.34 (0.13 to 0.56) | 0.002 |
| **Change in trend** | 0.26<br>(0.09 to 0.43) | 0.002 | 0.23<br>(0.07 to 0.40) | 0.006 | 0.00<br>(-0.01 to 0.01) | 0.790 |
| **R<sup>2</sup>** | 0.542 |  | 0.600 |  | 0.201 |  |

Although there was no statistically significant change in level, there was a statistically significant increasing trend in actual utilisation following the adoption of the tool (**Table 3**, **Figure 1B**). At six months, actual utilisation was 91.7%, compared with an expected 78.8% (95% CI: 73.0 to 84.6%) without the FDP Inpatient CCS. Mean actual utilisation for the whole post-intervention period was 12.2% (95% CI: 10.8 to 13.5%, p < 0.0001) higher than expected compared to the counterfactual estimate.

There was also a step-change increase in bookings per session after implementation of the FDP Inpatient CCS tool (**Table 3**, **Figure 1C**). By six months, the observed median was 3.8 bookings per session, compared with a modelled estimate of 3.5 (95% CI: 3.2 to 3.8) had the FDP Inpatient CCS not been introduced. Across the whole post-intervention study period, mean bookings per session was 10.9% (95% CI: 9.5 to 12.4%, p < 0.0001) higher than expected under the counterfactual scenario.

#### Utilisation by speciality

For the utilisation metrics, positive trend or level effects of the intervention were observed for Urology, General Surgery, Gynaecology, Plastic Surgery, and/or Ophthalmology (**Supplementary Appendix Table S1**). For the remaining specialities, although median weekly utilisation was higher on average post-intervention for many specialities (**Supplementary Appendix Table S2**), after accounting for pre-existing trends there were no statistically significant effects of the intervention detected.

#### Cancellations

After the introduction of the FDP Inpatient CCS, there was no significant level change in the percentage of cancelled cases (17.6; 95% CI: −10.8 to 55.0), p = 0.255), but the cancellation rate increased by an estimated 2.1% per week (95% CI: 0.9 to 3.2, p < 0.001) (**Figure 2**). At 6 months without the tool, the counterfactual model estimated a cancellation rate of 2.6% (95% CI: 1.8 to 3.8), compared to an observed cancellation rate of 4.9% after adopting the tool. For the whole post-intervention study period, 5.3% of bookings were cancelled, compared to an estimate 2.6% (95% CI: 1.8 to 3.8%) under the counterfactual estimate, reflecting that the mean cancellation rate was 98.2% (95% CI: 37.9 to 187.1%) higher compared to what was expected if the tool had not been adopted.

**Figure 2:**
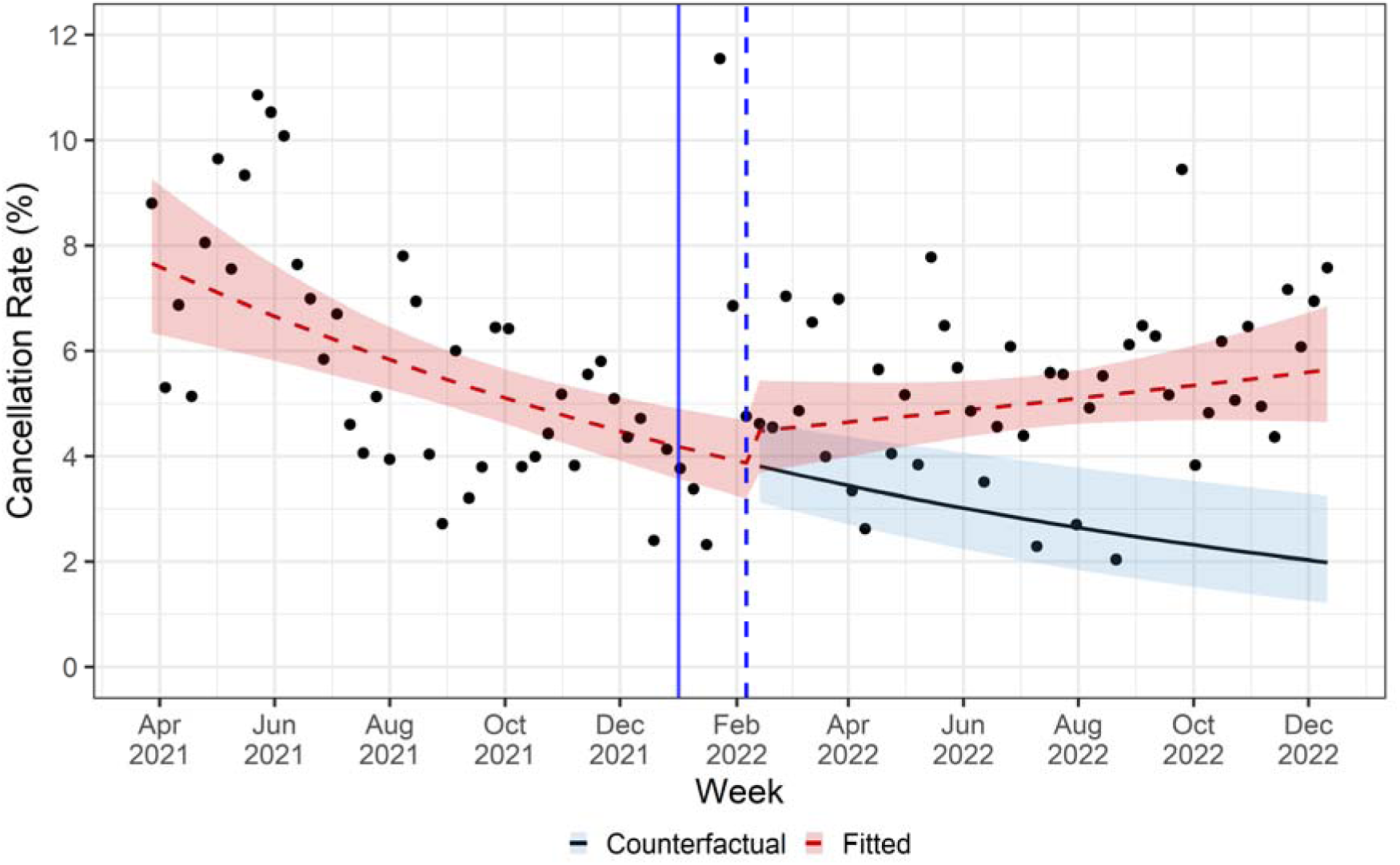
Changes in weekly rate of cancelled cases following introduction of the FDP Inpatient CCS product on January 3^rd^, 2022. The model assumes a five-week lag before the intervention takes effect, shown by the blue dashed line. The blue dashed line indicating the modelled five-week lag in the start-date. The red line shows the modelled fit to the observed results. The black line represents the counterfactual scenario (i.e., the expected results if there had been no intervention). The shaded areas around each line represent 95% confidence intervals.

## Discussion

### Summary of key findings

The implementation of the FDP Inpatient CCS product at CWFT in 2022 was linked to measurable improvements in theatre utilisation. Booked and actual utilisation were 15.0% and 12.2% higher, respectively, than counterfactual estimates. Additionally, the number of weekly bookings per session increased from 3.5 to 3.8 at six months, representing an overall 10.9% rise for the post-intervention period. Improvements in theatre utilisation metrics in Urology, General Surgery, Gynaecology, Plastic Surgery, and Ophthalmology specialty teams. Differences across specialities may relate to variation in implementation timing, workflow integration and service demand.

Although the post-intervention period was associated with a 2.1% weekly increase in cancellation rates (p < 0.001), the absolute change was negligible, with median cancellation rates remaining stable at 5.2% before and after implementation of the tool. The apparent rise in the trend of cancellations may partly reflect post-pandemic recovery effects and improved recording accuracy enabled by the new scheduling system, suggesting that while operational efficiency improved, data transparency and reporting mechanisms also evolved concurrently.

### Comparison to existing literature

Earlier systematic reviews have examined the role of machine learning and artificial intelligence in perioperative medicine, as well as theatre and theatre planning.[20,48–55] However, they did not focus on preoperative scheduling or the practical, real-world use of such algorithms. Consequently, the existing literature on digital scheduling tools is sparse, and therefore, our ability to conduct cross-study comparisons is limited.[56]

The few studies that examine similar scheduling tools do report improvements in performance and productivity.[57–59] All are observational, relying on descriptive or group-comparison statistics without modelling secular trends, which is an approach addressed by the ITS design employed in the current study. For example, the adoption of a digital scheduling system significantly reduced overall scheduling time [57], while a machine-learning-based tool that predicts case complexity produced more consistent scheduling results.[58] At Meyer Children’s Hospital, implementing a scheduling tool increased surgical throughput and reduced unused operating room time, despite a 12% capacity reduction due to external decisions.[59] This aligns with the present study’s findings, which showed an increase in bookings per theatre session and higher booked and actual utilisation, indicating performance improvements associated with the use of such tools. However, a pathway redesign intervention that incorporated a digital surgery scheduling tool reported a reduction in cancellations, which contrasts with our findings. As the intervention was multifaceted, it remains unclear whether the observed reduction can be attributed solely to the scheduling system.

### Strengths and Limitations

This study employed a quasi-experimental ITS design, a robust method commonly used in health service research, to evaluate the impact of the FDP Inpatient CCS.[45] This approach enabled comparison of trends before and after the intervention, capturing both level changes and gradual shifts over time. The ITS design, which accounted for underlying trends, was particularly relevant given the pre-intervention upward trend in utilisation, reflecting recovery from COVID-19-related declines in 2020 (**Supplementary Appendix Figure S11-13**). A major strength of this study is the large sample size, encompassing 31,572 surgical bookings, which enhances the statistical power of the findings. The study also integrated quality improvement methodology, combining programme theory with quantitative analyses to ensure methodological rigour and practical relevance.

However, several limitations should be considered. Two key modelling assumptions may have been violated: (I) that the intervention occurred at a single, discrete time point, and (II) that no external time-varying effects influenced outcomes. The implementation of the FDP Inpatient CCS tool was neither instantaneous nor uniform across all teams. The legacy scheduling system remained available throughout the study, allowing for variability in timing and extent of uptake. Internal promotion efforts, potentially linked to pandemic recovery plans, may also have contributed to the increased adoption patterns. Consequently, observed effects may not accurately reflect the full impact of the FDP Inpatient CCS theatre management platform, and its influence may be underestimated.

Time-varying external factors may also have affected theatre utilisation. Broader service initiatives and resource changes could have confounded the attribution of effects to the tool. Additionally, the limited 45-week pre-intervention period restricted full assessment of seasonal variation; although seasonality was partially accounted for using Fourier terms, residual effects may remain. This short pre-intervention timeframe also limits interpretation of longer-term impacts. Additionally, despite excluding data from the COVID-19 emergency period, the impact of pandemic-related disruptions may persist. For example, there is a decline in cancellations in 2021, which may reflect recovery from pandemic-related disruptions (**Supplementary Appendix Figure S13**), potentially introducing bias into the pre-intervention trends. Future analyses incorporating uptake patterns and adjustments for seasonal and external influences would provide a more precise understanding of the tools’ impact.

### Implications for research

This study contributes to the limited empirical evidence on the effectiveness of digital surgery scheduling tools.[56] Future research should employ robust designs, such as an ITS or randomised controlled trials, to strengthen causal inference and enable cross-study comparison. Given the brief pre-intervention period in this study, limited by COVID-19 service disruptions, longer assessment periods are necessary to determine if the observed improvements are sustained over time.

Further analyses could also explore the impact of these tools on referral-to-treatment performance and waiting list size, linking theatre efficiency to the recovery of elective care. Research should also examine implementation strategies that support consistent and scalable adoption. Understanding variation in use across teams (i.e., whether due to implementation factors or local context) would clarify how these digital scheduling tools are integrated into workflows. Qualitative research could explore the increasing cancellations to test whether the simplified cancellation process, enabled by a centralised tool, is responsible for this upward trend. Qualitative research could explore the increasing cancellations to test whether the simplified cancellation process, enabled by a centralised tool, is responsible for this upward trend.

### Implications for policy and practice

These findings demonstrate that digital platforms that integrate data across systems can enhance operational efficiency. Elective waiting lists remain a national priority, and recovery efforts increasingly focus on productivity improvement. National programmes such as the NHS *High Volume Low Complexity* and the *Getting It Right First Time* (GIRFT) have set a target of 85% theatre utilisation for Integrated Care by 2025.[60,61] Integrated digital technologies, such as the FDP, are consistently highlighted in NHS reform policies, including the NHS 10 Year Plan,[62] the Elective Care Recovery Plan,[23] and the Darzi Report,[63] as key enablers of productivity. However, prior to this study, no detailed evaluation of the tool’s operational impacts had been conducted.

This analysis confirms that centralising scheduling through platforms such as the FDP Inpatient CCS theatre management tool can enhance theatre capacity and reduce reliance on manual processes. This aligns with GIRFT priorities and helps prevent human errors. Enhanced scheduling efficiency benefits patients by minimising wait times and reducing the risks associated with delays. This strengthens the argument for adopting such tools to address the backlog in elective care. Sustained benefit, however, depends on consistent adoption across teams. To maximise gains, providers should ensure baseline engagement across all services, invest in staff training and embed digital tools into routine clinical workflows.

## Conclusion

The introduction of the FDP Inpatient CCS product for digital elective surgery scheduling at CWHT was linked to improved theatre utilisation, meeting GIRFT targets and suggesting a more efficient use of existing capacity. This study adds to the limited body of evidence on the effectiveness of digital scheduling systems for surgical services, which play a key role in the digital transformation of elective care. The findings may provide reassurance that centralised scheduling through such platforms can help theatres operate closer to full capacity. However, given the short pre-intervention period and the constraints of post–COVID-19 recovery, further research is needed to assess whether these improvements in utilisation are sustained over time and to examine variations in use across teams and specialties.

## Data Availability

Data are not publicly available. Data used in this study are derived from the Chelsea and Westminster Hospital NHS Foundation Trust via the NHS Federated Data Platform and were shared with Imperial College London for the purpose of this analysis. Data may be accessible through a separate agreement with Chelsea and Westminster Hospital NHS Foundation Trust.

## Ethics approval

This work was approved as a service evaluation by Chelsea and Westminster Hospital NHS Foundation Trust (Reference No. QI599).

## Transparency statement

As the manuscript’s guarantors, EL and GK affirm that the manuscript is an honest, accurate, and transparent account of the study and no important aspects of the study have been omitted.

## Contributor statement

ALN contributed to the conception of the study. EL and GK accessed and verified the data. GK and EL conducted data cleaning, analysis and presentation. EL and GK wrote the manuscript and accept full responsibility for the work and decision to publish, as its guarantors. NB and CH facilitated the project through enabling data access and assisted with interpretation of findings in the context of Trust operations. All authors provided critical revision and approved the final version of the manuscript.

## Copyright/licence for publication

The Corresponding Author has the right to grant on behalf of all authors and does grant on behalf of all authors, a worldwide licence to the Publishers and its licensees in perpetuity, in all forms, formats and media (whether known now or created in the future), to i) publish, reproduce, distribute, display and store the Contribution, ii) translate the Contribution into other languages, create adaptations, reprints, include within collections and create summaries, extracts and/or, abstracts of the Contribution, iii) create any other derivative work(s) based on the Contribution, iv) to exploit all subsidiary rights in the Contribution, v) the inclusion of electronic links from the Contribution to third party material where-ever it may be located; and, vi) licence any third party to do any or all of the above.

