## Supplementary materials for "Impact of the Federated Data Platform’s digital surgery scheduling system on elective theatre utilisation at an NHS Trust: an interrupted time series analysis"

| **CONTENTS** | **Page:** |
| --- | --- |
| **Tool overview** | **2** |
| Supplementary Figure S1 | 3 |
| Supplementary Figure S2 | 4 |
| Supplementary Figure S3 | 5 |
| Supplementary Figure S4 | 6 |
| Supplementary Figure S5 | 7 |
| **Logic Model** | **8** |
| Supplementary Figure S6 | 8 |
| **Model Development** | 9 |
| Supplementary Figure S7 | 10 |
| Supplementary Figure S8 | 11 |
| Supplementary Figure S9 | 12 |
| Supplementary Figure S10 | 12 |
| **Supplementary Tables** | **13** |
| Supplementary Table S1 | 13 |
| Supplementary Table S2 | 14 |
| Supplementary Table S3 | 15 |
| **Supplementary Figures** | **16** |
| Supplementary Figure S11 | 16 |
| Supplementary Figure S12 | 16 |
| Supplementary Figure S13 | 17 |

**Tool Overview**

The NHS Federated Data Platform (FDP) IP theatre management tool is an FDP IP product used to manage elective theatre scheduling. Figure 1 is a snapshot of the tool from the user’s perspective. The tool provides a unified user interface to view and edit theatre session information. Supplementary Figures 2-5 provide overviews of the steps a user may take to execute a desired action.

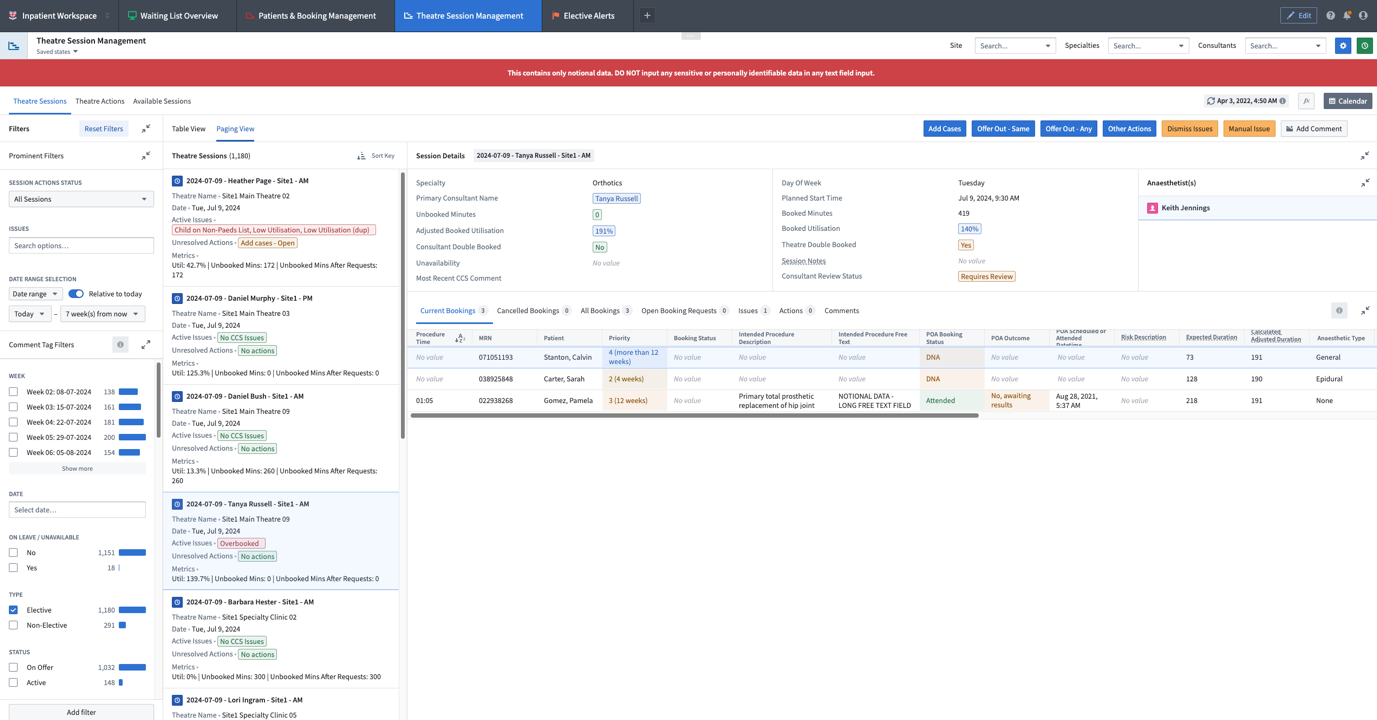

**Supplementary Figure S1:** Screenshot of the NHS FDP IP Theatre Session Management module with dummy data.

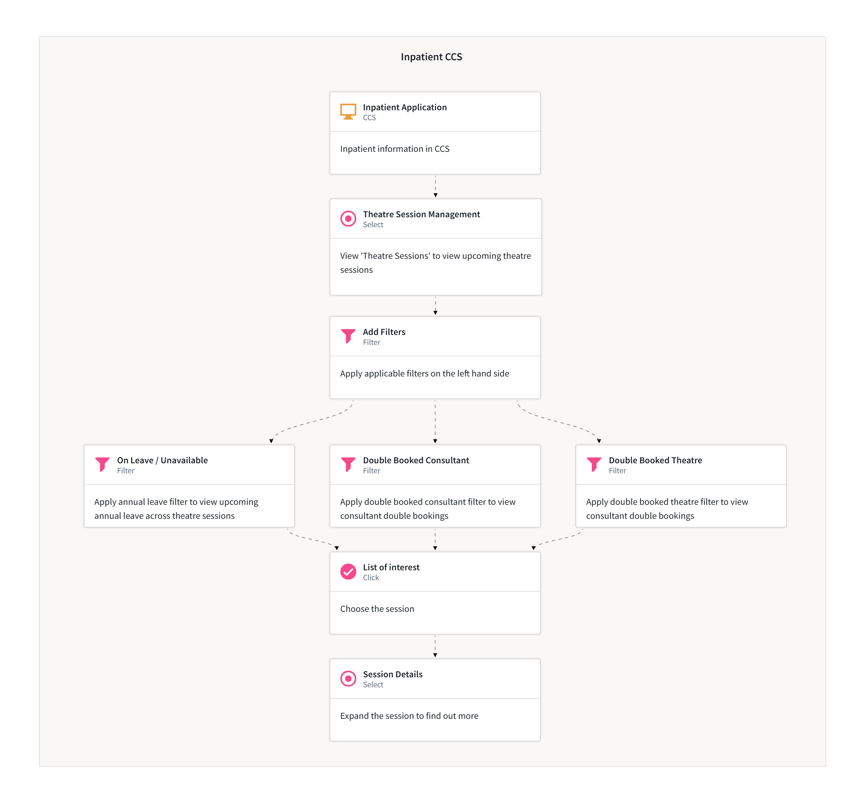

**Supplementary Figure S2**: Flowchart demonstrating how users may use the FDP IP theatre management module to view and offering out available theatre sessions.

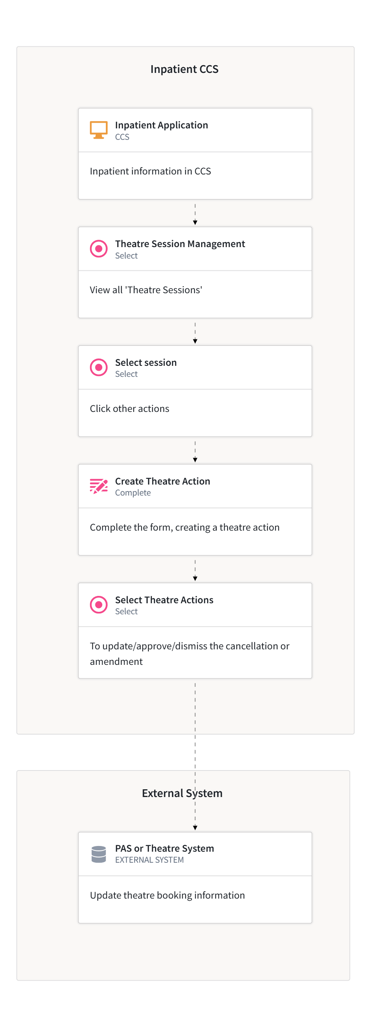

**Supplementary Figure S3:** Flowchart demonstrating how users may use the FDP IP theatre management module to cancel a case or amend a theatre session.

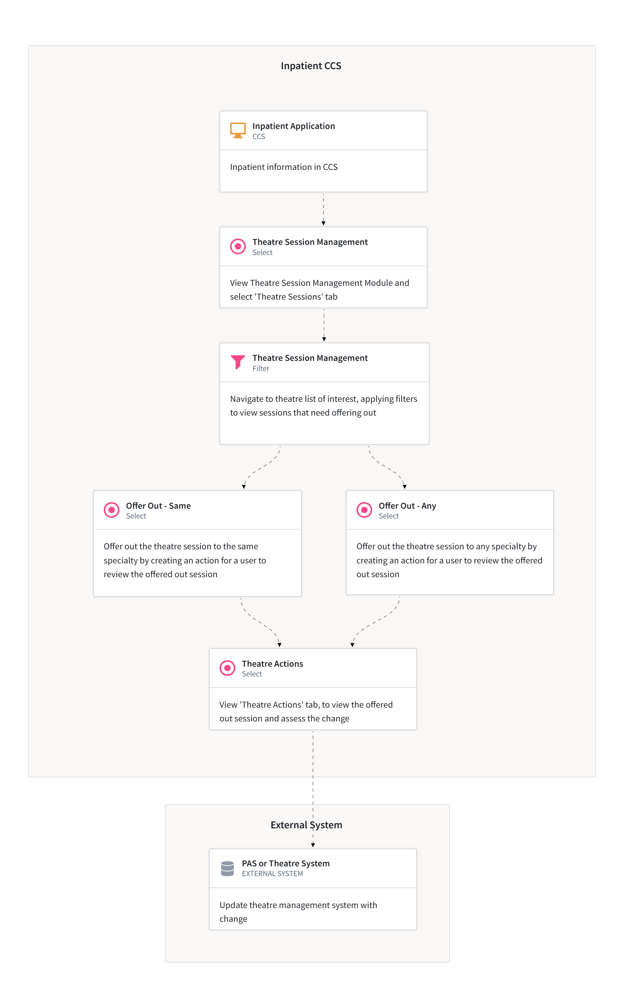

**Supplementary Figure S4:** Flowchart demonstrating how users may use the FDP IP theatre management module to check theatre sessions for issues and amendments.

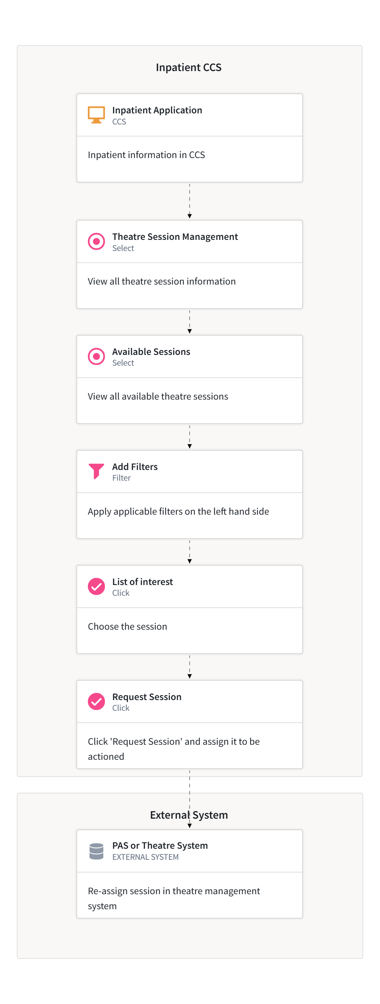

**Supplementary Figure S5:** Flowchart demonstrating how users may use the FDP IP theatre management module to view and request a theatre session.

**Logic Model**

To identify appropriate metrics for the evaluation, a two-hour hybrid workshop was convened on 19 July 2024, prior to the commencement of quantitative analysis. The aim was to develop a shared understanding of the FDP IP platform, particularly its theatre session management features, drawing on the expertise of subject matter specialists and related workflows. Seven participants attended, including subject matter experts from delivery teams across three NHS trusts (THH, ICHT, and LNWUH). The research team acted as facilitators and observers, taking detailed notes, recording and transcribing the session, and conducting a thematic analysis to identify key concepts and inform evaluation metrics. Complementary sources included online training materials for the inpatient module and standard operating procedures (SOPs) from three NHS trusts, incorporating workflow maps and stakeholder-specific guides (e.g. for managers, schedulers, clinicians, and theatre staff). These materials provided further insight into platform use across contexts and informed the development of a programme theory, which depicts the causal pathways between activities and outcomes using visual formats such as logic models or driver diagrams (**Supplementary Figure 6**).

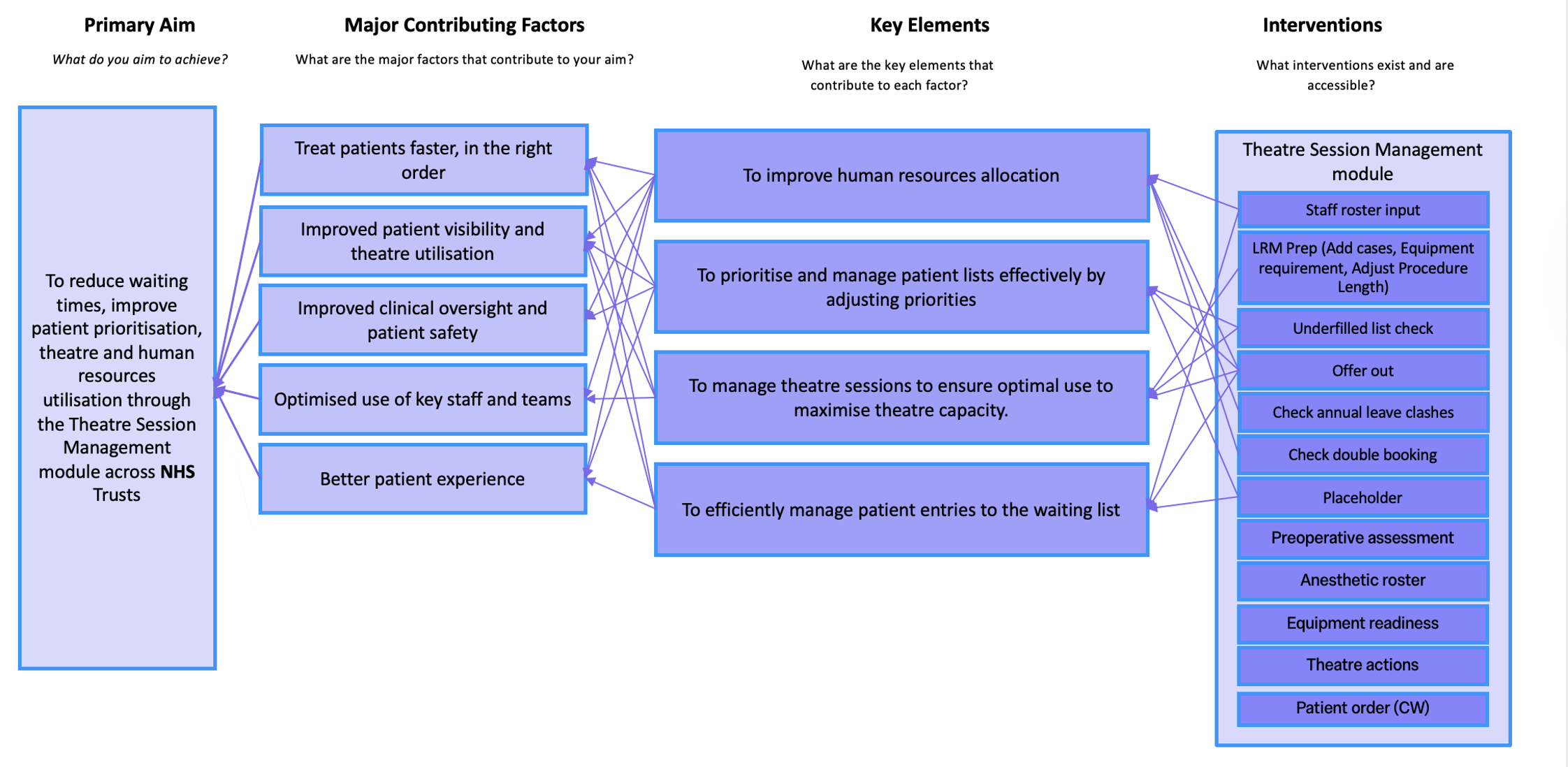

**Supplementary Figure S6:** Logic model for the FDP IP, theatre management module

**Model Development**

Data were accessed via the FDP secure environment. Elective theatre sessions for obstetric or paediatric services were excluded.

*Autocorrelation*

The weekly utilisation (booked utilisation, actual utilisation, bookings per session [BPS]) and cancellation metrics were first visually inspected using histograms, Autocorrelation Function (ACF) and Partial Autocorrelation Function (PACF) plots.

The utilisation metrics were first modelled using ordinary least squares (OLS) segmented regression and the residuals inspected for evidence of autocorrelation. For BPS and cancellations, the residuals for OLS models showed no evidence of residual autocorrelation and the final models were therefore OLS models with no autoregressive structures. For booked and actual utilisation, OLS models exhibited evidence of autoregression of order 1 (AR(1)), meaning each value depended on the previous value. Therefore, instead of OLS models, generalised least squares (GLS) models were used with an AR(1) structure for the booked and actual utilisation models.

*Seasonality*

Visual inspection suggested seasonal fluctuations in utilisation metrics which should be accounted for in regression models. Fourier terms are pairs of sine and cosine functions used to model periodic (cyclical) patterns in time series. They capture smooth, repeating seasonal patterns without assuming a specific functional form. These sine and cosine pairs are included in the models as covariates, allowing the model to adjust the mean outcome for seasonal cycles.

$$Fourier terms of order k: \left\{ \begin{aligned} \sin(\frac{2\pi kt}{S}) \\ cos(\frac{2\pi kt}{S}) \end{aligned} \right.$$

Where:

- $t$ = 1, 2, …, $n$ = time index
- $S$ *=* length of the seasonal cycle (i.e., the number of observations per period)
- $k$ = 1, 2, …, $K$ = number of harmonics, where a higher $k$ captures higher frequency fluctuations

For each outcome, the selection of $S$ and $k$ was achieved through grid searching combinations of their values and selecting the model with the lowest Bayesian Information Criteria (BIC). The search ranges were 2 to 52 for $S$ and 1 to 8 for $k$.

For actual and booked utilisation, the optimal values for $S$ and $k$ were 37 and 2, respectively, while for BPS, the optimal values were 35 and 1, respectively. These values are unexpected given weekly data would be expected to exhibit a periodicity of 52; however, this data is limited to 90 weeks total, or 45 weeks in each segment, meaning the data structure may inhibit the ability to fully capture seasonal effects.

Fourier terms were not found to improve the model of cancellation rates; consequently, no seasonal components were ultimately included in the cancellation rate models.

For the specialty-level analyses, the same process, as described above, was performed for checking autocorrelation and Fourier terms.

*Model diagnostics*

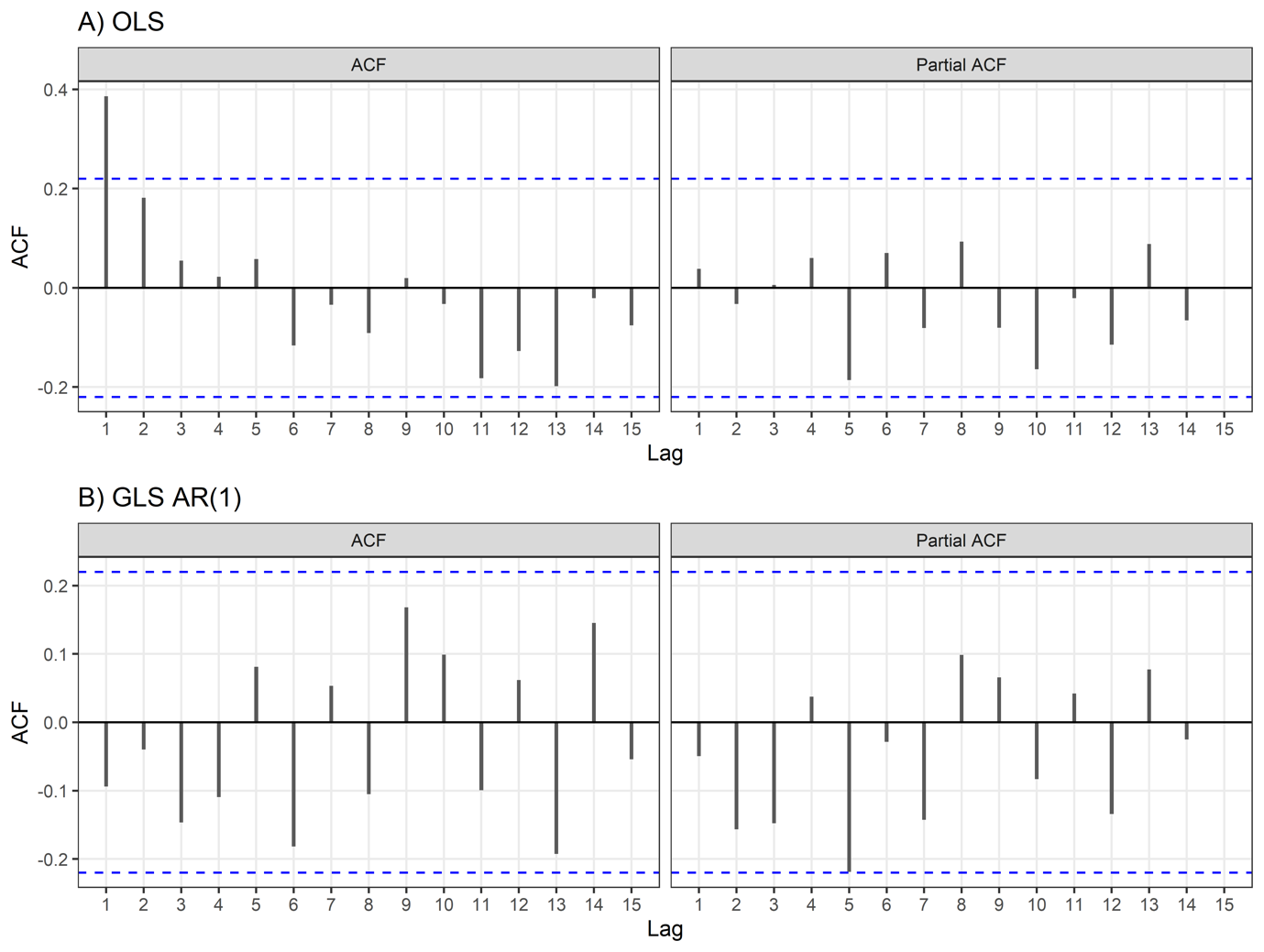

**Supplementary Figure S7: ACF and PACF plots of residuals from the A) initial OLS regression model and B) final GLS regression model with an AR(1) structure and Fourier terms, for models of weekly median booked utilisation.** Residuals for the final model followed a normal distribution (Shapiro-Wilk test: *P*=835), were homoscedastic (Breusch-Pagan test: *P* =0.476), and were stationary (KPSS test: *P>*0.100). The Autocorrelation Function plots and Ljung-Box test up to lag 15 (*P*=0.124) suggest there is no remaining autocorrelation in the residuals.

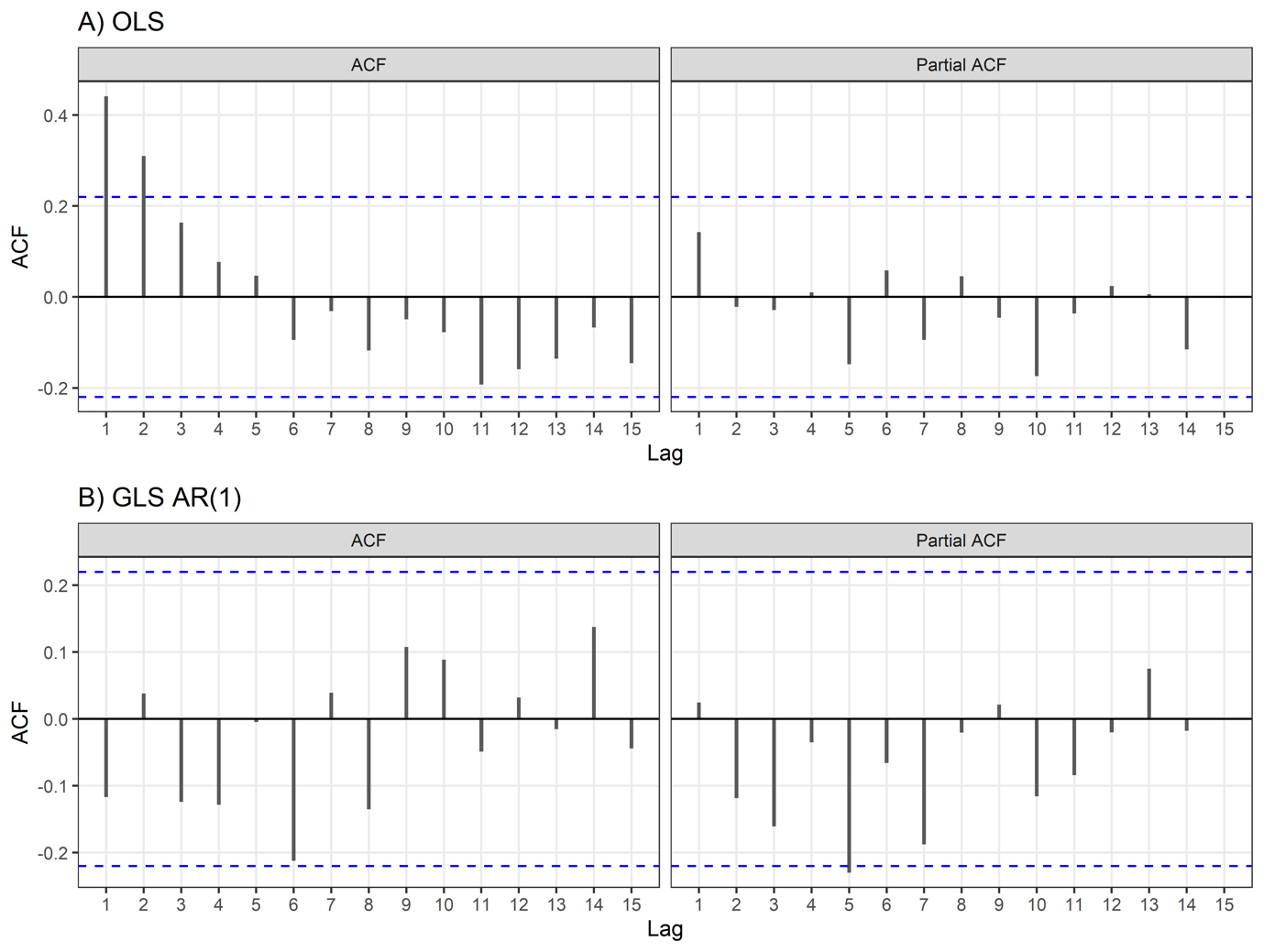

**Supplementary Figure S8: ACF and PACF plots of residuals from the A) initial OLS regression model and B) final GLS regression model with an AR(1) structure and Fourier terms, for models of weekly median actual utilisation.** Residuals for the final model followed a normal distribution (Shapiro-Wilk test: *P*=0.129), were homoscedastic (Breusch-Pagan test: *P* =0.222), and were stationary (KPSS test: *P>*0.100). The Autocorrelation Function plots and Ljung-Box test up to lag 15 (*P*=0.412) suggest there is no remaining autocorrelation in the residuals.

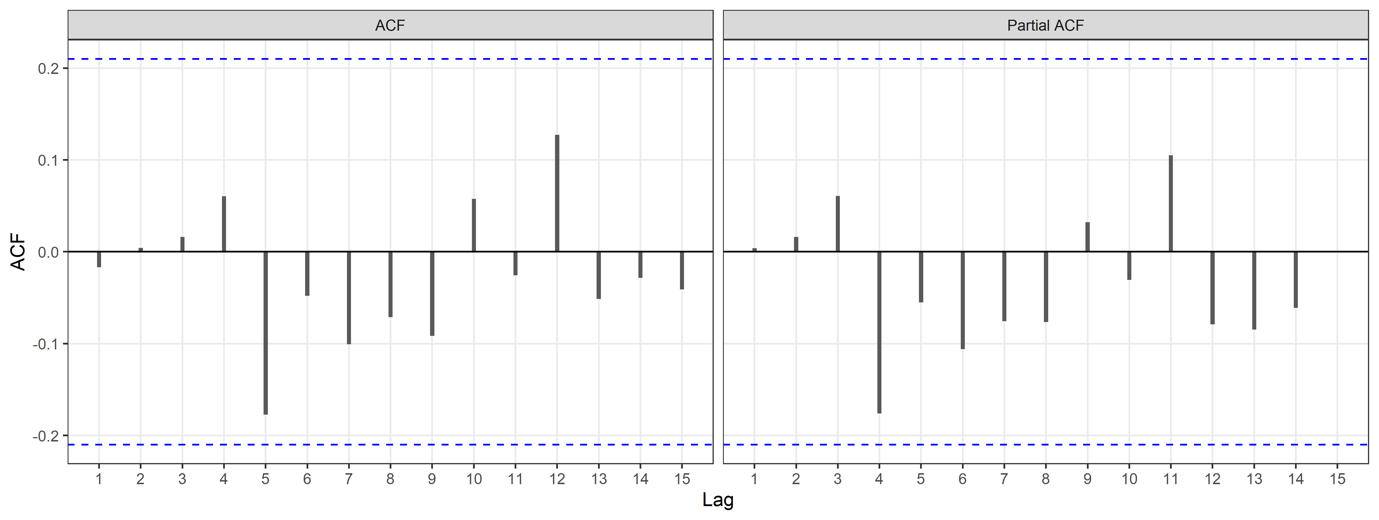

**Supplementary Figure S9: ACF and PACF plots of residuals from the final OLS regression model of weekly median number of bookings per session with Fourier terms.** Residuals for the final model followed a normal distribution (Shapiro-Wilk test: *P*=0.380), were homoscedastic (Breusch-Pagan test: *P* =0.462), and were stationary (KPSS test: *P>*0.100). The Autocorrelation Function plots and Ljung-Box test up to lag 15 (*P*=0.846) suggest there is no remaining autocorrelation in the residuals.

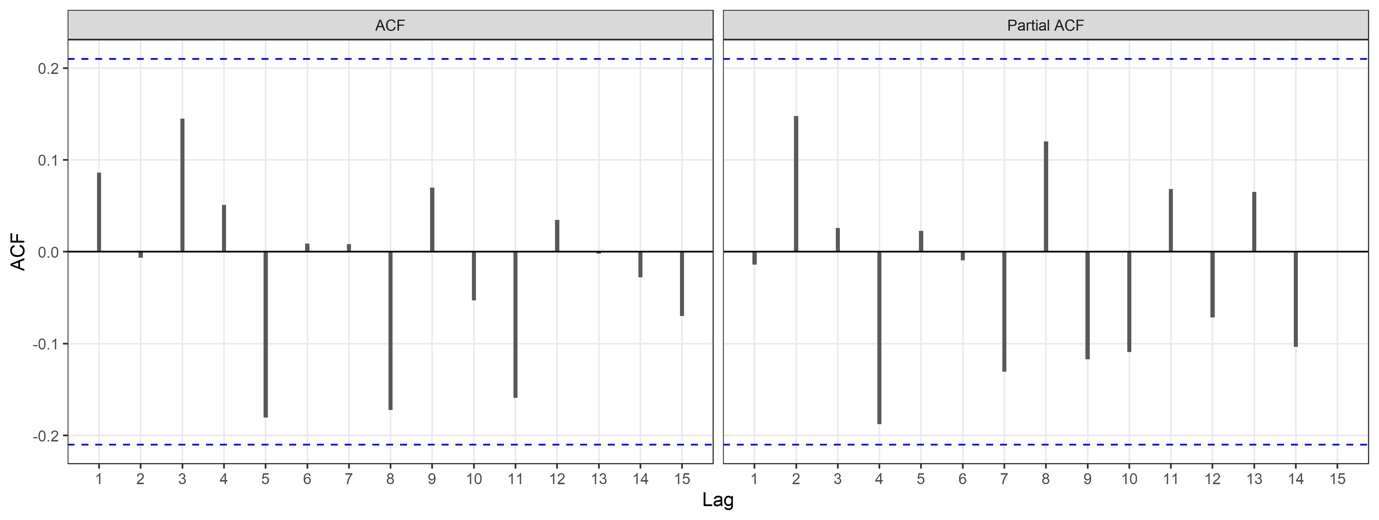

**Supplementary Figure S10: ACF and PACF plots of residuals from the final OLS regression model of weekly percentage of cancelled bookings.** Residuals for the final model followed a normal distribution (Shapiro-Wilk test: *P*=0.138), were homoscedastic (Breusch-Pagan test: *P* =0.141), and were stationary (KPSS test: *P>*0.100). The Autocorrelation Function plots and Ljung-Box test up to lag 15 (*P*=0.501) suggest there is no remaining autocorrelation in the residuals.

**Supplementary Tables**

**Supplementary Table S2: Results from interrupted time series analyses of median weekly booked and actual utilisation and weekly bookings per session. The order column describes the autoregressive (p) and/or moving average (q) terms employed in the model.**

|  | **Booked Utilisation** | | | | **Actual Utilisation** | | | | **Bookings per theatre session** | | | |
| --- | --- | --- | --- | --- | --- | --- | --- | --- | --- | --- | --- | --- |
|  | **β (95% CI)** | **p-value** | **Order** | **Fourier** | **β (95% CI)** | **p-value** | **Order** | **Fourier** | **β (95% CI)** | **p-value** | **order** | **Fourier** |
| **Urology** |  |  | None | S=11, K=1 |  |  | None | None |  |  | None | None |
| Level impact | 6.21  (-0.01 to 12.4) | 0.053 |  |  | 9.61  (2.86 to 16.36) | 0.001 |  |  | 7.05 (0.58 to 13.53) | 0.033 |  |  |
| Change in trend | 0.01  (-0.22 to 0.25) | 0.919 |  |  | 0.09  (-0.17 to 0.35) | 0.493 |  |  | 0  (-0.25 to 0.25) | 0.982 |  |  |
| **General Surgery** |  |  | None | S=45, K=1 |  |  | None | S=42, K=1 |  |  | None | None |
| Level impact | 9.22  (1.05 to 17.4) | 0.03 |  |  | 5.73  (-1.81 to 13.3) | 0.14 |  |  | 0.21  (-0.10 to 0.52) | 0.182 |  |  |
| Change in trend | 0.00  (-0.22 to 0.21) | 0.976 |  |  | 0.02  (-0.21 to 0.25) | 0.864 |  |  | 0.02  (0.01 to 0.03) | 0.002 |  |  |
| **Trauma and Orthopaedics** |  |  | None | S=12, K=1 |  |  | None | None |  |  | None | None |
| Level impact | -3.72  (-10.00 to 2.56) | 0.249 |  |  | -4.31  (-10.3 to 1.69) | 0.163 |  |  | 0.01 | 0.935 |  |  |
| Change in trend | 0.15  (-0.09 to 0.39) | 0.221 |  |  | 0.13  (-0.11 to 0.36) | 0.293 |  |  | 0 .00  (-0.01 to 0.02) | 0.419 |  |  |
| **Gynaecology** |  |  | p=1, q=0 | S=43, K=1 |  |  | p=1, q=0 | S=37, K=1 |  |  | None | None |
| Level impact | 12.60  (4.03 to 21.1) | 0.005 |  |  | 7.90  (1.40 to 14.4) | 0.02 |  |  | 0.16  (-0.21 to 0.52) | 0.395 |  |  |
| Change in trend | 0.44  (0.16 to 0.72) | 0.003 |  |  | 0.52  (0.26 to 0.78) | <0.001 |  |  | 0.01 (-0.01 to 0.02) | 0.431 |  |  |
| **Plastic Surgery** |  |  | None | S=40, K=6 |  |  | p=1, q=0 | S=41, K=6 |  |  | p=1, q=1 | None |
| Level impact | 31.6  (17.8 to 45.5) | <0.0001 |  |  | 22.8  (4.49 to 41.00) | 0.017 |  |  | 0.38  (0.02 to 0.74) | 0.038 |  |  |
| Change in trend | 0.27  (-0.21 to 0.75) | 0.276 |  |  | 0.26  (-0.37 to 0.89) | 0.417 |  |  | 0.01  (0.00 to 0.03) | 0.064 |  |  |
| **Colorectal Surgery Service** |  |  | None | None |  |  | None | None |  |  | p=1, q=0 | 8,1 |
| Level impact | -5.39  (-14.3 to 3.49) | 0.237 |  |  | -4.58  (-14.7 to 5.54) | 0.378 |  |  | 0.38  (-0.23 to 0.98) | 0.218 |  |  |
| Change in trend | 0.21  (-0.14 to 0.55) | 0.242 |  |  | 0.23  (-0.16 to 0.62) | 0.25 |  |  | 0.01  (-0.01 to 0.04) | 0.244 |  |  |
| **Ophthalmology** |  |  | None | S=15, K=7 |  |  | p=1, q=0 | S=15, K=7 |  |  | None | 10,1 |
| Level impact | 4.92  (-5.42 to 15.3) | 0.354 |  |  | 5.96  (-4.43 to 16.3) | 0.265 |  |  | 0.97  (0.00 to 1.93) | 0.049 |  |  |
| Change in trend | 0.68  (0.3 to 1.06) | <0.0001 |  |  | 0.90  (0.51 to 1.29) | <0.0001 |  |  | 0.08  (0.04 to 0.11) | <0.0001 |  |  |

**Supplementary Table S1:** Median utilisation and interquartile range (IQR), comparing pre- and post- intervention using Wilcoxon tests.

| **Specialty** | **Pre-intervention,**  **Median (IQR)** | **Post-intervention,**  **Median (IQR)** | **P-value** |
| --- | --- | --- | --- |
| *Booked Utilisation* |  |  |  |
| Urology | 63.0 (57.7 to 66.7) | 62.5 (58.3 to 66.7) | 0.728 |
| General Surgery | 76.4 (72.2 to 81.2) | 84.3 (78.5 to 88.9) | 0.001 |
| Trauma and Orthopaedics | 79.7 (74.1 to 83.3) | 79.2 (75.2 to 86.7) | 0.244 |
| Gynaecology | 79.4 (71.9 to 83.7) | 87.9 (80.9 to 91.7) | <0.0001 |
| Plastic Surgery | 66.7 (55.3 to 81.1) | 85.2 (77.5 to 89.3) | <0.0001 |
| Colorectal Surgery Service | 77.8 (74.1 to 84.7) | 85.2 (70.4 to 88.9) | 0.134 |
| Ophthalmology | 71.7 (60.7 to 78.8) | 75.0 (70.2 to 86.5) | 0.023 |
| *Actual Utilisation* |  |  |  |
| Urology | 68.8 (62.5 to 75.0) | 67.7 (62.5 to 74.0) | 0.792 |
| General Surgery | 85.6 (79.7 to 90.6) | 90.6 (87.5 to 97.4) | 0.002 |
| Trauma and Orthopaedics | 85.1 (80.0 to 89.8) | 87.5 (83.3 to 91.7) | 0.115 |
| Gynaecology | 81.2 (75.0 to 87.5) | 91.7 (84.8 to 93.8) | <0.0001 |
| Plastic Surgery | 71.2 (58.0 to 83.8) | 86.7 (81.2 to 93.3) | <0.0001 |
| Colorectal Surgery Service | 87.5 (80.7 to 93.8) | 93.8 979.2 to 100.0) | 0.127 |
| Ophthalmology | 75.0 (64.2 to 82.5) | 77.3 (72.1 to 87.9) | 0.07 |

**Supplementary Figure Table S3**: Percentage of bookings which were cancelled by specialty, comparing pre- and post- intervention using Chi-Squared tests.

| **Specialty** | **Pre-intervention** | **Post-intervention** | **Statistic** | **P-value** |
| --- | --- | --- | --- | --- |
| Urology | 5.98 (4.00 – 9.00) | 3.80 (3.39 – 4.41) | 1374.5 | 0.0035 |
| General Surgery | 5.23 (4.00 – 6.15) | 5.51 (4.00 – 6.67) | 975.5 | 0.771 |
| Trauma and Orthopaedics | 6.12 (4.65 – 7.69) | 7.26 (5.49– 9.50) | 846 | 0.239 |
| Gynaecology | 4.55 (3.92 – 6.20) | 3.51 (2.30 – 4.29) | 1364 | 0.00454 |
| Plastic Surgery | 0.00 (0.00-2.70) | 4.17 (0 – 5.72) | 631 | 0.00717 |
| Colorectal Surgery Service | 0.00 (0.00 – 5.26) | 5.57 (0.00 – 7.69) | 791 | 0.115 |
| Ophthalmology | 0.00 (0.00 – 5.56) | 4.00 (0.00 – 8.33)) | 877.5 | 0.245 |

**Supplementary Figures**

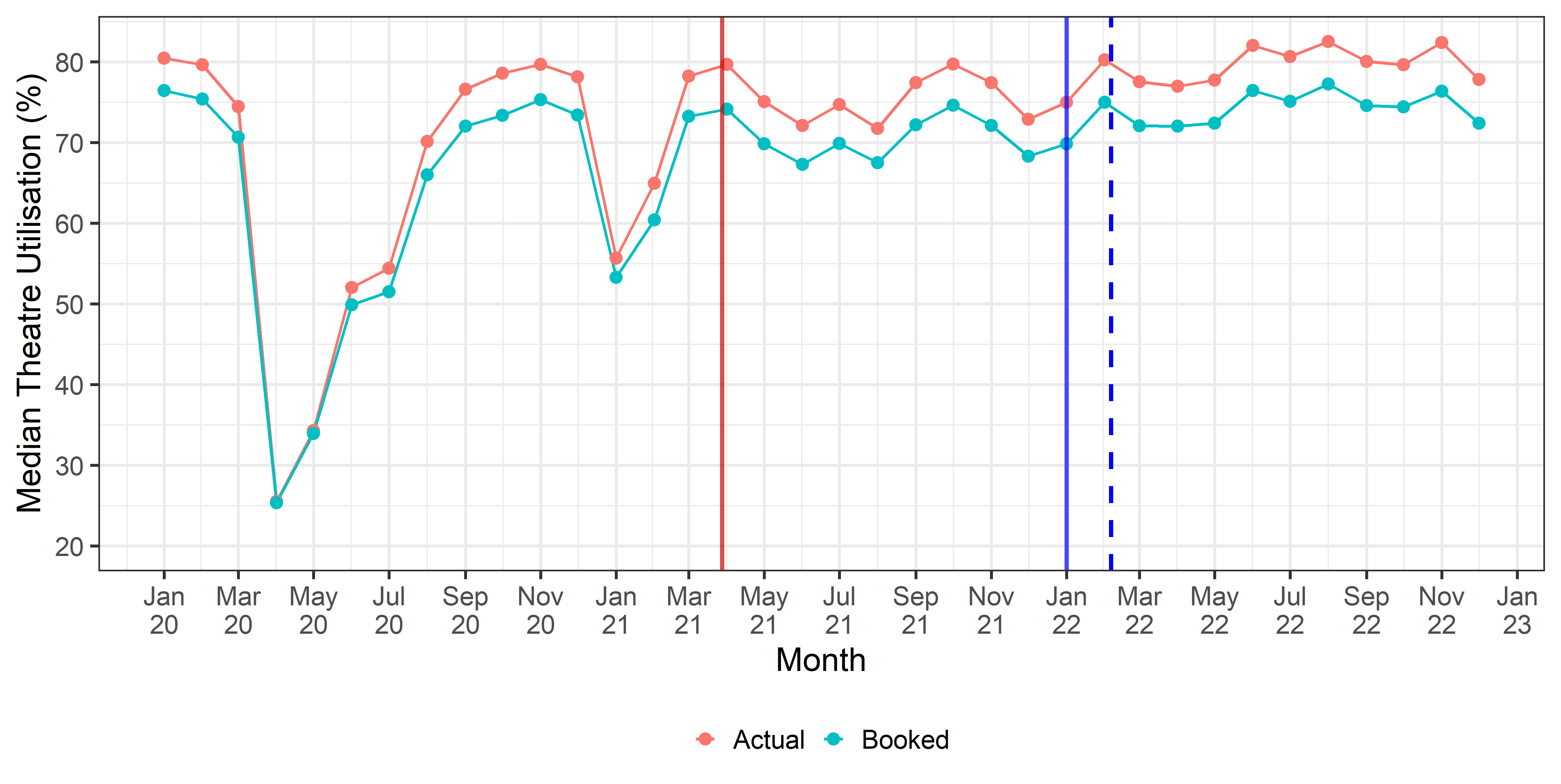

**Supplementary Figure S11:** Median booked and actual theatre utilisation over time. The red line represents the start of the study period, while the solid blue line marks the date of FDP IP adoption. The dashed blue line indicates the modelled adoption date when incorporating the 5-week lag.

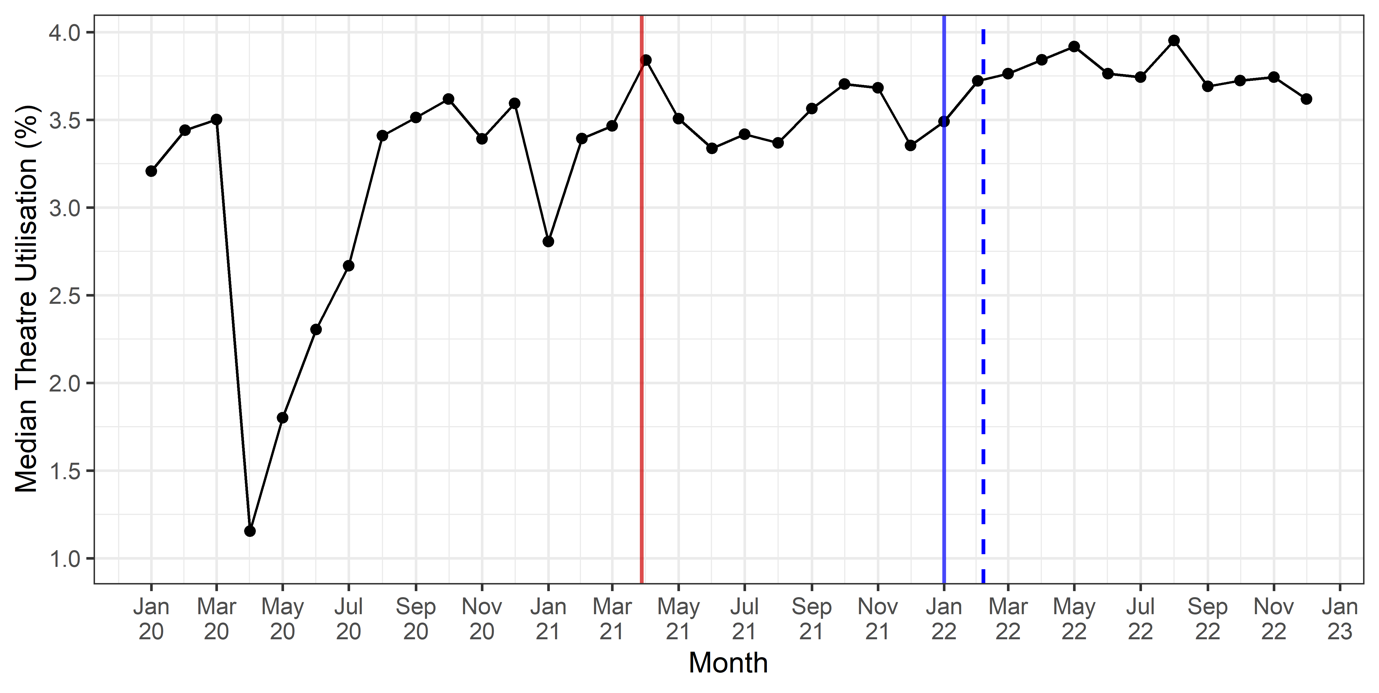

**Supplementary Figure S12:** Median booked and actual theatre utilisation over time. The dashed red line represents the start of the study period, while the solid blue line marks the date of FDP IP adoption. The dashed blue line indicates the modelled adoption date when incorporating the 5-week lag.

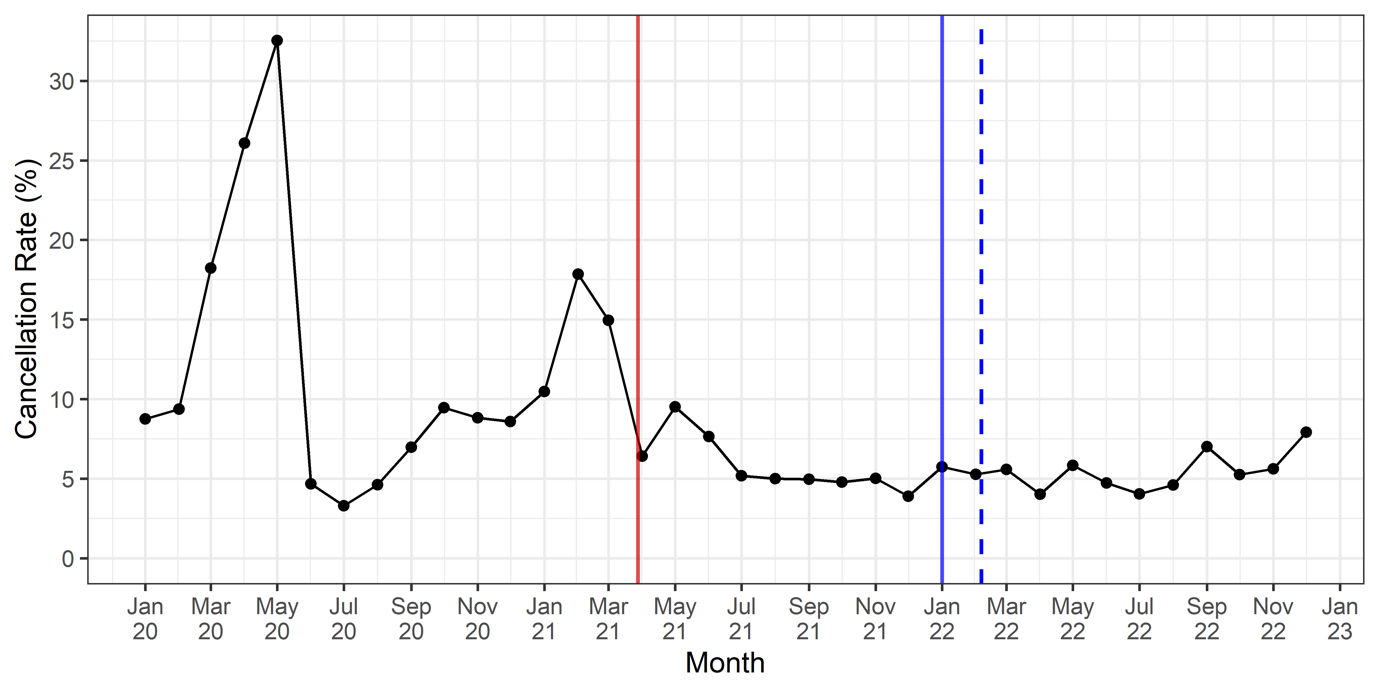

**Supplementary Figure S13:** The percentage of cancelled bookings by month. The dashed red line represents the start of the study period, while the solid blue line marks the date of FDP IP adoption. The dashed blue line indicates the modelled adoption date when incorporating the 5-week lag.
